# Ambient environmental exposures and mental health outcomes in Mozambique: a population-based nationally representative survey

**DOI:** 10.1101/2025.10.29.25339102

**Authors:** Geoffrey Barini, Walter Mogeni, Polycarp Mogeni

## Abstract

**Background:** Although the impact of climate change on mental health is widely acknowledged, there is a critical lack of empirical evidence from sub-Saharan Africa. We used a nationally representative cross-sectional study conducted in Mozambique to assess whether ambient environmental exposures are associated with symptoms of anxiety or depression.

**Methods:** Mental health data from the 2022-2023 Mozambique Demographic and Health Survey were linked to remotely sensed climate and environmental variables, and associations were determined using the modified Poisson regression.

**Results:** A total of 13,138 women and 5,380 men were included in the study. The prevalence of anxiety was 11.1% among women and 2.1% among men, whilst depressive symptoms were 10.1% in women and 2.3% in men. Among women, higher mean daily temperatures (>24.6°C) were associated with an increased likelihood of symptoms of anxiety (adjusted risk ratio [aRR] = 2.89, 95% CI: 2.13-3.95; p < 0.0001) and depression (aRR = 3.45, 95% CI: 2.46-4.84; p < 0.0001). High annual precipitation (>1201mm) was associated with a >2-fold higher likelihood of either outcome, while relative humidity (68.4%-74.2%) was associated with a >2-fold higher likelihood of either outcome. In contrast, high environmental greenness (NDVI >0.56) was associated with at least a 49% decrease in symptoms of either outcome. No clear associations were observed among men.

**Implications:** Exposure to high temperatures, humidity, and precipitation was associated with an increased risk of adverse mental health outcomes among women in Mozambique. These findings underscore the need for targeted policy responses, including the development of green spaces and enhanced flood mitigation, to promote resilience and safeguard mental health among vulnerable populations.

**Research in Context:** *Evidence before this study:* There is limited evidence from sub-Saharan Africa on the mental health impacts of climate-related exposures such as temperature, humidity, precipitation, and vegetation cover. Most existing studies are from high-income or Asian settings, with few studies examining gender or age-specific vulnerabilities. Literature suggests that women may be disproportionately affected by climate stressors due to biological, social, and economic factors, though evidence from sub-Saharan Africa remains scarce. Age differences in climate-related mental health impacts are also understudied.

*Added value of this study:* To our knowledge, this is one of the first studies from Mozambique, and more broadly from Africa, to examine associations between climate factors and mental health outcomes. We found that higher temperatures, relative humidity, and precipitation were associated with a greater likelihood of depression, anxiety, and their co-occurrence. In contrast, higher environmental greenness was protective across these outcomes. Importantly, these associations were observed among women but not among men, highlighting a potential gender differential in vulnerability to environmental exposures.

*Implications of all the available evidence:* Our findings emphasize the urgent need to expand research on climate and mental health in African contexts, with attention to gender and age differences. The apparent greater vulnerability of women suggests that climate-related mental health impacts may exacerbate existing gender inequities. Interventions aimed at climate adaptation—including expanding green spaces and flood mitigation—could help protect mental health, particularly for women in similar settings.

## Introduction

Mental health disorders are a leading cause of disability worldwide, accounting for substantial morbidity and health care costs^1^. The Global Burden of Disease study ranks depression, anxiety, and post-traumatic stress disorder among the top contributors to years lived with disability, with the economic toll extending to workforce productivity and health system capacity^1^. While research has traditionally focused on psychosocial^2^, economic, and geopolitical stressors, genetics, and neurobiological mechanisms^3^, a growing body of evidence now highlights the critical role of environmental stressors related to climate change^4^.

Rising global temperatures and increasingly frequent and severe weather events, like heat waves, floods, and cyclones, pose unprecedented health risks^5^. Heat waves, for example, have been linked to more hospitalizations for psychiatric conditions, while floods and cyclones are associated with higher rates of PTSD, depression, and anxiety^5–7^. The mechanisms behind these links are both direct and indirect. Direct effects include physiological stress from heat, which can disrupt sleep and impair cognitive function, as well as neuroinflammatory responses from prolonged thermal exposure^8^. Indirectly, climate events amplify mental health risks through pathways such as displacement, economic insecurity, and food system disruption, though the full extent of these impacts remains unquantified^7^.

The evidence base is especially scarce for sub-Saharan Africa, where mental health infrastructure is underdeveloped and climate vulnerability is high^9^. Mozambique exemplifies this dual burden: recurrent cyclones, erratic rainfall, and progressive warming threaten both public health and livelihoods, with cascading consequences for psychological well-being^10–13^. Recent demographic surveys indicate high rates of depression, anxiety, and PTSD^14–16^, yet few studies have examined the contribution of environmental exposures to this burden.

Here, we aimed to investigate the relationship between ambient environmental factors and mental health outcomes in Mozambique. We analyze nationally representative data from Mozambique to test the hypothesis that high temperatures, elevated humidity, and excessive precipitation worsen mental health outcomes, whilst environmental features such as green space may confer resilience. We integrated epidemiologic, climatic, and demographic data to assess the potential environmental determinants of mental illness in this vulnerable setting and identify potential preventive interventions.

## Methods

### Study setting

This secondary analysis was conducted using data from the 2022-23 Mozambique demographic and health survey (MDHS)^17^. The Demographic and Health Surveys (DHS) are nationally representative surveys conducted by government agencies in low and middle-income countries (LMIC) with support from the U.S Agency for International Development (USAID). The DHS uses standardized questionnaires and sampling methods to collect detailed information on country-specific demographics and health data, including mental health, and socioeconomic indicators, among others^18^.

### Study design and data collection

The 2022–23 MDHS was a two-stage, stratified cluster sampling design to generate nationally representative estimates as well as disaggregated estimates by urban–rural residence, province (including Maputo City), and major geographic regions (coastal, border, and inland). In the first stage, 619 enumeration areas (EAs) were selected from the 2017 census frame using probability proportional to size, stratified by urban–rural location; 232 EAs were urban and 387 were rural. Eight districts in Cabo Delgado were excluded due to security concerns. In the second stage, a household listing exercise was conducted in each EA to create an updated sampling frame, from which 26 households were systematically selected, yielding a final sample of 16,045 households after the exclusion of two additional EAs for security reasons. Interviews were carried out with all women aged 15–49 residing in the selected households. For men, eligibility was restricted to those aged 15–54 residing in a randomly selected 50% of households. A detailed description of the sampling procedures and criteria for participation in the mental health modules has been published elsewhere^19^.

### Exposure variables

Environmental and climatic exposure variables included: (a) total annual precipitation, (b) mean daily air temperature (i.e., air temperature measured at 2 m above the ground surface, representing near-surface conditions), (c) mean daily relative humidity, (d) normalized difference vegetation index (NDVI), (e) mean daytime air temperature and relative humidity, (f) mean nighttime air temperature, (g) mean nighttime relative humidity, and (h) the Universal Thermal Climate Index (UTCI). NDVI was obtained from the Moderate Resolution-Imaging Spectro-Radiometer (MODIS) sensor aboard NASA’s Terra satellite^20^, as provided by the NASA Land Processes Distributed Active Archive Center; precipitation data were obtained from Climate Hazard Group InfraRed Precipitation Station^21^; while mean air temperatures and relative humidity products were obtained from the European Center for Medium-Range Weather Forecasts ERA5-Land reanalysis^22^. ERA5 has been rigorously validated and is widely recognized as a reliable data source for studies on temperature-related health impacts^23^. Since relative humidity (%) is not directly available in ERA5-Land products, it was derived using humidity-related variables, which include air temperature and dew point temperature (appendix p1). Data on thermal comfort were obtained from GLoUTCI^24^. All the environmental variables were resampled using nearest interpolation and exported as raster files at 500m spatial resolution through Google Earth Engine’s JavaScript API^25^. These environmental variables were extracted at the DHS survey cluster level by calculating the mean value within a 5 km buffer for rural clusters and a 2 km buffer for urban clusters, reflecting the standard spatial displacement applied to protect survey respondents’ confidentiality^26^. Hourly ambient temperature and relative humidity, and the normalized difference vegetation index (16-day composite) were averaged over the 12 months preceding the 2022-2023 MDHS survey to capture chronic exposure, while total annual precipitation during the same period was used to capture cumulative rainfall conditions.

### Outcome variables

Our primary outcome variables were: a) symptoms of generalized anxiety, b) depressive symptoms, and c) concurrent symptoms of anxiety and depression. Symptoms of generalized anxiety and symptoms of depression were assessed using the Generalized Anxiety Disorder scale (GAD-7) and the Patient Health Questionnaire (PHQ-9), respectively. GAD-7 and PHQ-9 have been rigorously validated and recognized as effective screening instruments for identifying individuals at increased risk of mental disorders in both primary care settings and the general population^27,28^.

For anxiety, respondents were asked how often they had experienced any of the symptoms of anxiety two weeks preceding the survey, with scores ranging from 0 (never) to 3 (always), and the total score was used to assess the severity of anxiety symptoms. Similarly, to assess depressive symptoms, respondents reported the frequency with which they had experienced the symptoms of depression two weeks before the survey, with scores ranging from 0 (never) to 3 (always), and a total score of 0 to 27 used to assess the severity of depressive symptoms.

For both disorders, we used a cut-off score of ≥10 to define the presence of clinically significant symptoms of anxiety or depression^29,30^. Respondents with a prior diagnosis of anxiety or depression from a doctor, and who were taking medication for either condition, were classified as positive for symptoms of anxiety or depression, regardless of their GAD-7 or PHQ-9 scores.

### Statistical analysis

Sex-stratified analyses were performed, with women and men analyzed separately to assess potential differences in associations by sex. Climatic and environmental variables were categorized into three groups—low (<25th percentile), moderate (25th–75th percentile), and high (>75th percentile)—to account for potential nonlinear relationships with mental health outcomes. Associations between climatic and environmental exposures and mental health outcomes were examined using modified Poisson regression models.

Sensitivity analyses were conducted to evaluate the robustness of the findings. First, daily 2 m air temperature and relative humidity were replaced with corresponding daytime and nighttime measures, modeled separately to avoid collinearity and to assess potential differences in their psychological and epidemiological effects. Second, the Universal Thermal Climate Index (UTCI) was used as an alternative composite measure of thermal stress. Subgroup analyses stratified by age among women (15–22, 23–32, and 33–49 years) were conducted to assess potential differences in associations across age groups.

All models were adjusted for potential confounders, including age (15–19, 20–24, 25–29, 30–34, 35–39, 40–44, and 45–49 years for women; with an additional 50–54 category for men), education level (none, primary, or secondary/higher), wealth index (quintiles), marital status (never married, married, or divorced/separated), self-rated health (poor, moderate, or good), and residence (urban or rural). These covariates were selected based on established associations with mental health outcomes and their potential to confound the relationships of interest. For categorical variables with more than two levels, we used global Wald tests to assess overall associations.

Adjusted risk ratios (aRR) and the 95% confidence interval (95%CI) were estimated using the modified Poisson regression. Statistical analyses were two-sided, with p-values < 0.05 deemed statistically significant, and were performed using R software version 4.3.3 (https://www.r-project.org/). All statistical analyses accounted for the complex survey design.

**Fig 1.**
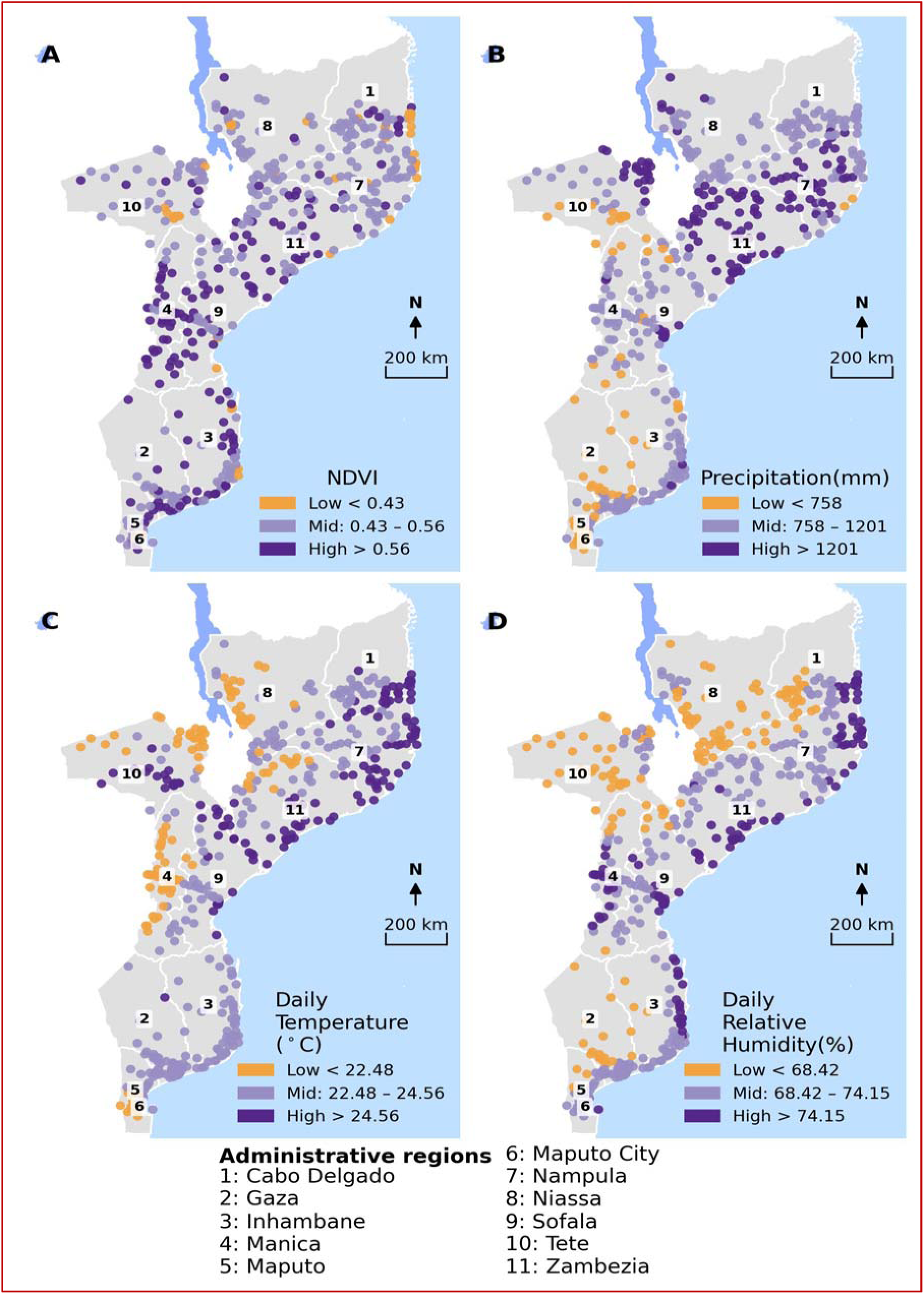
Spatial distribution of environmental factors at DHS survey locations. (A): Normalized Difference Vegetation Index, (B): Total annual precipitation, (C): 2m air temperature, (D): Relative humidity.

## Results

### Characteristics of study participants

The study sample included 13,183 women aged 15–49 years and 5,380 men aged 15–54 years. Among these, 30.8% of women and 41.7% of men had attained at least secondary education. Employment levels differed markedly by sex, with 69.7% of women not employed compared with 18.8% of men. More than twice as many women as men were divorced or separated (appendix p. 1). Self-reported severe poor health was less common among women (1.1%) than among men (5.2%).

### Spatial distribution of climatic and environmental factors

The mean daily air temperature and daily relative humidity in the study area ranged from 18.4 °C to 26.3 °C, with a median of 23.8 °C (IQR 22.5 - 24.9 °C), and daily relative humidity ranged from 52.6% to 78.5%, with a median of 72.3% (IQR 69.3% −74.3%). Although daytime and nighttime relative humidity variables followed similar spatial patterns (appendix p4), median daytime relative humidity was substantially lower than nighttime relative humidity (63.4% vs. 85.2%) (appendix p4). Additionally, the spatial patterns of nighttime temperature and relative humidity were consistent with those observed during daytime (appendix p4). Vegetation health, as indicated by NDVI was moderate with a median of 0.51 (IQR 0.42-0.55) (appendix p4). The total annual precipitation recorded across the survey clusters ranged from 159mm to 1999mm, with a median of 989mm (IQR 758-1201).

### Prevalence of mental health symptomology and ambient environment risk factors

The prevalence of mental health symptoms was substantially higher among women than men, including anxiety (11.1% vs. 2.1%), depression (10.1% vs. 2.3%), and co-occurring symptoms (7.3% vs. 0.7%), respectively. Among women, prevalence rates were marked with geographical variation, with the highest burden observed in Nampula and Zambézia provinces (Figure 2). Furthermore, symptoms of anxiety, depression, and co-occurrence among women—but not men—increased with higher ambient temperature exposure (anxiety: from 5.2% in relatively low ambient temperature regions to 18.5% in relatively high ambient temperature areas, Ptrend=0.003; depression: 3.8% to 17.4%, Ptrend=0.001; co-occurrence: 2.5% to 12.9%, Ptrend=0.004; appendix p). Similar patterns were observed for precipitation (appendix p). A graded association was observed between temperature and mental health outcomes among women in multivariable analyses. Compared with those exposed to temperatures below the 25th percentile, women in the moderate range (25th–75th percentile) had a 45% higher risk of anxiety symptoms (aRR = 1.45; 95% CI, 1.06–1.97; p = 0.0196), whereas those exposed to high temperatures (>75th percentile; >24.6 °C) had nearly a threefold higher risk (aRR = 2.89; 95% CI, 2.13–3.93; p < 0.0001). A similar graded pattern was evident for depressive symptoms, with women in the moderate range associated with a 79% higher risk (aRR = 1.79; 95% CI, 1.26–2.56; p = 0.0012) and those residing in the high-temperature range associated with more than a threefold higher risk (aRR = 3.45; 95% CI, 2.46–4.84; p < 0.0001). This pattern was consistent for the co-occurrence of anxiety and depression (Figure 3). Higher relative humidity levels were significantly associated with an increased risk of anxiety, depression, and their co-occurrence (Wald test, 2 df; all p < 0.05) (Figure 3, appendix p. 4). Compared with women residing in areas receiving precipitation below the 25th percentile, those in the highest precipitation category (>75th percentile) had a higher risk of anxiety (aRR = 1.98; 95% CI, 1.35–2.92; p = 0.0005), depression (aRR = 2.20; 95% CI, 1.41–3.41; p = 0.0005), and co-occurring symptoms (aRR = 2.09; 95% CI, 1.20–3.64; p = 0.0098). In contrast, residents of greener environments (NDVI >0.56; >75th percentile) were associated with at least 49% lower likelihood of anxiety, depression, and co-occurring symptoms (Figure 3). Among men, no clear associations were observed between most environmental exposures and mental health outcomes, except for relative humidity, which was unexpectedly associated with a lower likelihood of depressive symptoms (Figure 4).

**Fig 2.**
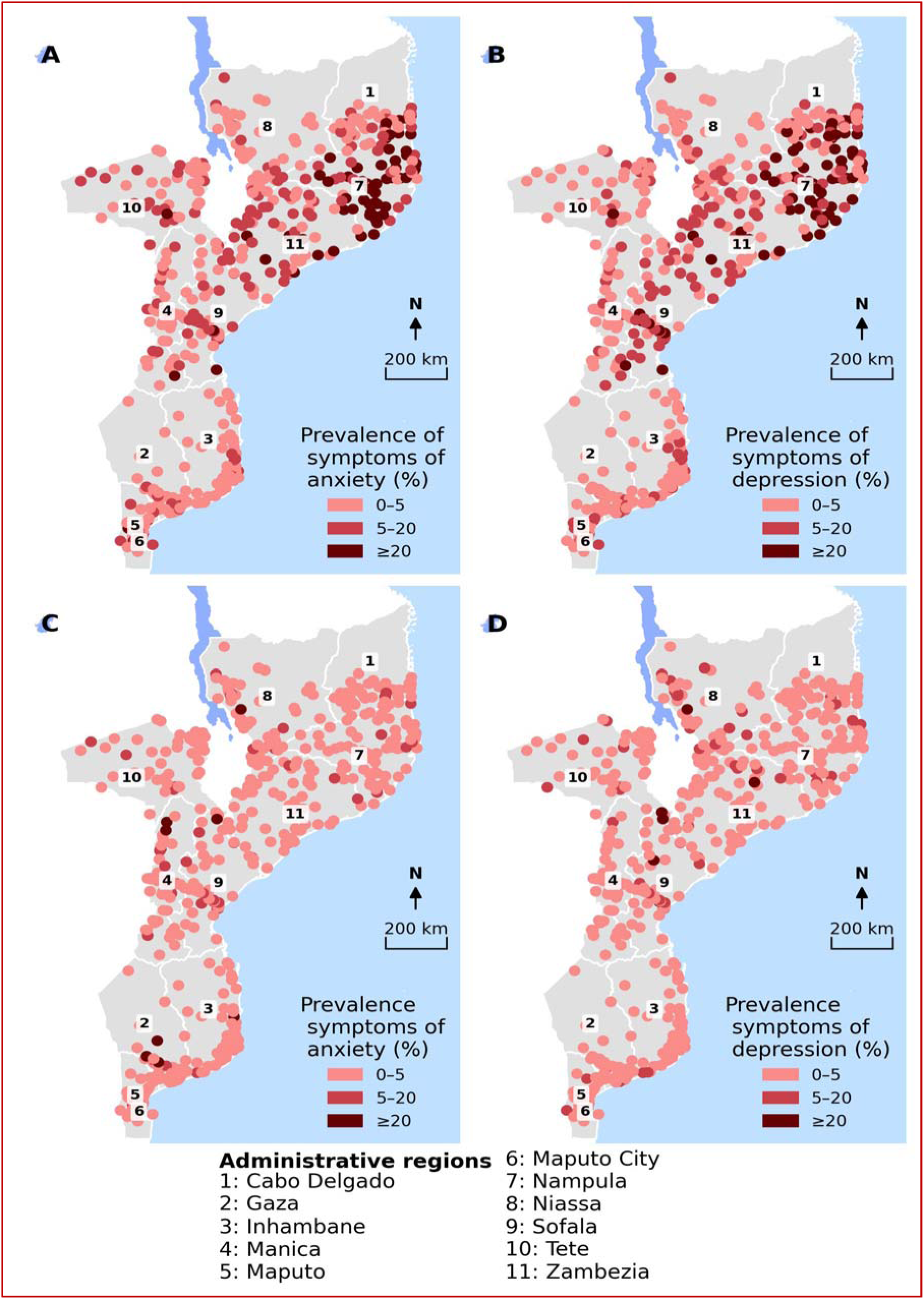
Map showing spatial distribution of survey weighted prevalence estimates of symptoms of anxiety and depression by DHS survey locations. Panels (A) and (B) for women and Panels (C) and (D) for men.

**Figure 3.**
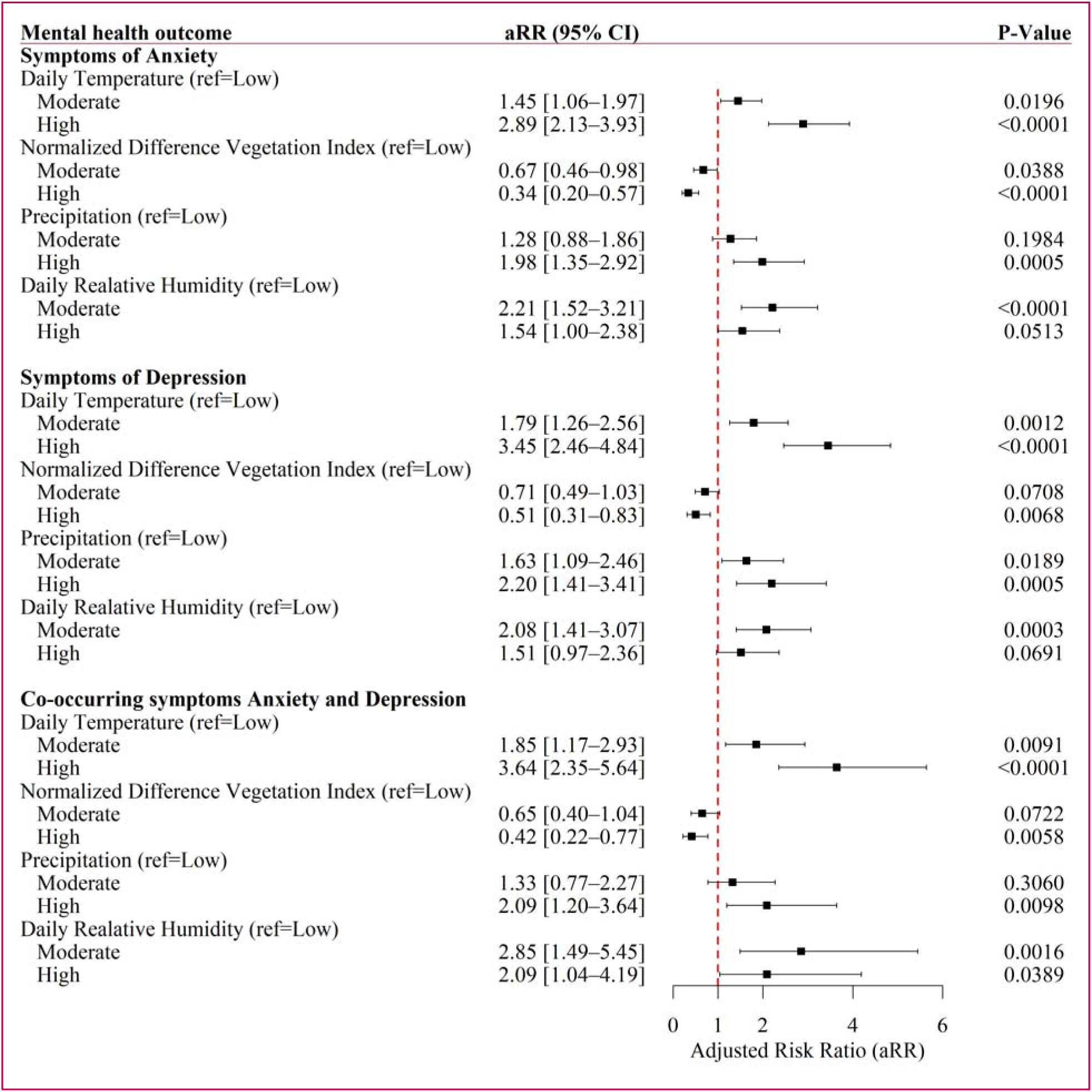
Estimates of adjusted risk ratios of the associations between ambient environmental factors and mental disorders among women in Mozambique. The models were adjusted for age, education, health status, urbanity, marital status, employment and household wealth quintile. The categories low (<25the percentile), moderate (25th-75th percentile) and high (>75the percentile) reflect increasing exposure burden for environmental factors.

**Figure 4.**
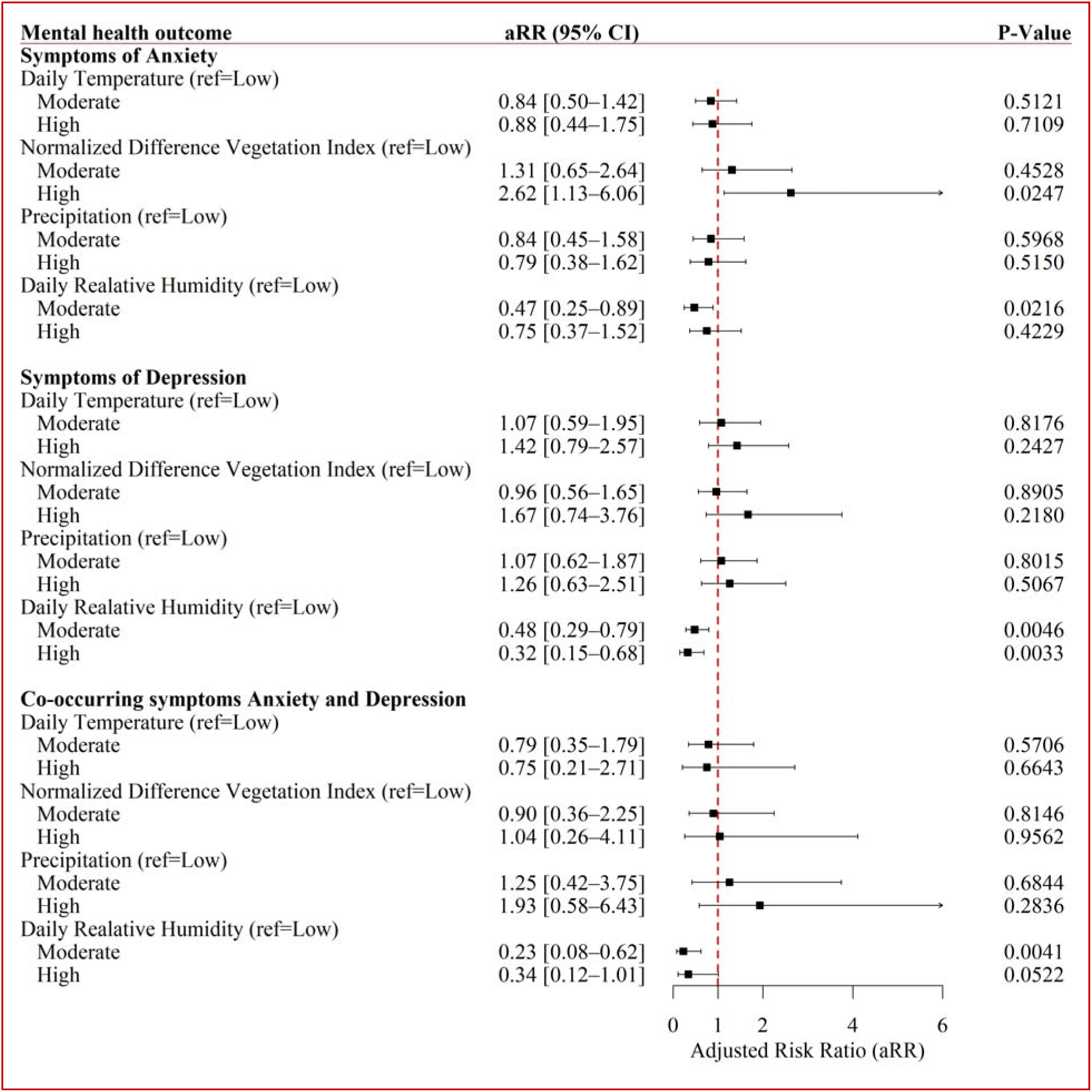
Estimates of adjusted risk ratios of the associations between and ambient environmental factors and mental disorders among men in Mozambique. The models were adjusted for age, education, health status, urbanity, marital status, employment and household wealth quintile. The categories low (<25the percentile), moderate (25th-75th percentile) and high (>75the percentile) reflect increasing exposure burden for environmental factors

In sensitivity analyses that examined daytime and nighttime temperatures separately among women, the associations remained consistent but were more pronounced for daytime temperature, whereas those with nighttime temperature were weaker (appendix p 8). In contrast, the associations between relative humidity and mental health outcomes differed by time of day. Nighttime relative humidity was consistently and significantly associated with higher likelihood of all mental health outcomes, while daytime humidity showed weaker and mostly nonsignificant associations (appendix p 8). Higher UTCI values, reflecting greater thermal stress derived from the combined effects of temperature, humidity, wind speed, and radiation, were associated with an increased likelihood of adverse mental health outcomes, consistent with the main findings (appendix p 8).

In subgroup analyses, the adverse effects of daily temperature on mental health outcomes among women were consistent across all age groups, whereas associations with relative humidity varied by age (appendix p 9). In contrast, the protective associations of greener environments were evident only among women aged 33–49 years, while high precipitation was consistently associated with poorer mental health outcomes within this age group (appendix p 9).

## Discussion

In this secondary analysis of nationally representative data from Mozambique, we examined the associations between ambient environmental conditions and common mental health outcomes, including anxiety, depression, and their co-occurrence. Women residing in geographical areas characterized by higher ambient temperatures, greater relative humidity, and higher precipitation had an increased likelihood of experiencing symptoms of common mental health disorders, whereas women residing in greener environments were associated with lower risks. In contrast, these associations were not observed among men. Together, these findings underscore the potential mental health burden associated with environmental stressors in tropical, low-income settings and demonstrate important gender differences in vulnerability.

Consistent with previous studies,^31–33^ higher ambient temperatures were associated with an increased likelihood of depressive and anxiety symptoms among women. Multiple pathways may explain this relationship, including physiological stress responses, psychological strain, and social or behavioral disruptions. Daytime and nighttime temperatures may influence mental health through different mechanisms: high daytime temperatures can reduce outdoor activity and social interaction—both important for psychological well-being—whereas elevated nighttime temperatures may interfere with sleep quality, contributing to anxiety and depressive symptoms^6,6,34^. These findings have important implications for LMIC such as Mozambique, where average temperatures are predicted to increase by approximately 0.3 °C per decade between 2000 and 2050^12^.

The association between ambient relative humidity and adverse mental health outcomes is consistent with prior evidence linking humid conditions to worse psychological well-being and suicide risk. Epidemiological studies report positive associations between higher humidity and mental-health outcomes, with some analyses suggesting adverse effects among women and younger people^35^. Humid conditions may impair thermoregulation and sleep, increase physiological stress responses, and limit social and physical activity—factors that have been associated with the onset and exacerbation of anxiety and depressive symptoms in animal models^36^. A recent systematic review of temperature-related mental health studies highlights the importance of humidity and its interaction with temperature in predicting mental health risks, demonstrating considerable heterogeneity across populations and settings^37^. These studies support our finding of a humidity–mental health association and are compatible with the sex-specific pattern we observed. However, the evidence on sex differences remains limited and inconsistent across contexts, underscoring the need for further research in low-income and tropical settings^37^.

Higher annual precipitation was associated with poorer mental health outcomes among women, consistent with evidence from other regions^5^. In this context, high precipitation may serve as a proxy for periods of excessive rainfall or localized flooding, which can disrupt mobility, damage homes and crops, and intensify household and economic stress^10,38^. Such conditions often reduce social interaction, heighten caregiving burdens, and increase exposure to environmental and socioeconomic stressors—all of which are associated with anxiety and depressive symptoms. The disproportionate effects observed among women may reflect their greater responsibilities for caregiving, food provision, and household management in the aftermath of rainfall-related disruptions^33,39^.

Conversely, residence in greener environments, as reflected by higher NDVI values, was associated with a lower likelihood of anxiety, depression, and their co-occurrence. Our findings were consistent with prior research demonstrating that exposure to vegetation may promote psychological restoration, reduce stress, and strengthen social cohesion and physical activity^40,41^. Together, these findings suggest that while high temperature and precipitation may exacerbate environmental and social vulnerabilities affecting women, greenness may buffer against adverse mental health challenges^40^.

This study had some limitations. First, our analysis relied on secondary data that may be subject to social desirability and recall bias^42^. Given the stigma surrounding mental health, especially among men, underreporting may have occurred, potentially influencing both the estimated prevalence and the observed associations^43^. Averaging environmental exposures over a period of 12 months preceding the 2022-23 MDHS survey may obscure short-term or seasonal variations that may influence mental health outcomes. However, evidence from studies in tropical regions remains mixed, with some studies showing minimal seasonal variations in mental health outcomes, likely due to relatively stable temperature and daylight patterns through the year^44^. Furthermore, because the data are cross-sectional, it was not possible to establish the temporal sequence of mental health symptoms, limiting causal inference. Our differential findings between nighttime and daytime measures should be interpreted with caution, as ambient temperature and relative humidity were assessed at the cluster rather than the household or indoor level. Although nighttime measurements may partially correlate with indoor conditions, variations in housing structure and development across households could lead to differential exposure, potentially attenuating the observed associations with mental health outcomes. Nonetheless, this study is the first to examine the association between ambient environmental factors and mental health outcomes in Mozambique using nationally representative data. The analysis also controlled for a range of socioeconomic and demographic variables to mitigate potential confounding.

In conclusion, our analysis demonstrates that women in Mozambique bear a disproportionate climate-associated mental health risk, underscoring the vulnerability of women to climatic extremes coupled with gendered exposure patterns^32,33,45^. The observed differences between women and men in Mozambique underscore the urgent need to integrate the gender lens into climate-related mental health research and policy. Public health interventions addressing climate-driven mental health risks in LMICs should prioritize women as a high-risk population, with targeted interventions such as heat-related awareness, disaster recovery programs, and integration of mental health care into climate adaptation policies. Our findings contribute to growing evidence that climate change is not only an ecological crisis but also a driver of socioeconomic and mental health inequities. Mental health policymakers should promote coordinated strategies that integrate mental health, climate adaptation, and gender equity to address potential impacts of climate change on vulnerable populations.

## Authors’ contributions

GB and PM conceptualized and designed the study. GB curated data. GB, and PM conducted the literature review. GB and PM contributed to the data analysis. GB, WM, and PM contributed to the interpretation of the results. GB drafted the initial manuscript. GB, WM, and PM edited and approved the final manuscript. PM provided overall supervision.

**All authors have read and confirmed that they meet ICMJE criteria for authorship.**

## Data availability

The DHS data used in this study is available at https://dhsprogram.com/data/available-datasets.cfm and can be freely accessed by registered users.

