## Supplementary Appendix for "Ambient environmental exposures and mental health outcomes in Mozambique: a population-based nationally representative survey"

| **Characteristic** | **Category** | **Women**  **N (%)** | **Men**  **N (%)** |
| --- | --- | --- | --- |
| **Age group** | 15-19 | 3050 (23.1%) | 1386 (25.8%) |
|  | 20-24 | 2693 (20.4%) | 976 (18.1%) |
|  | 25-29 | 2195 (16.7%) | 781 (14.5%) |
|  | 30-34 | 1577 (12%) | 635 (11.8%) |
|  | 35-39 | 1486 (11.3%) | 500 (9.3%) |
|  | 40-44 | 1171 (8.9%) | 446 (8.3%) |
|  | 45-49 | 1011 (7.7%) | 390 (7.2%) |
|  | 50-54 | - | 266 (4.9%) |
| **Residence** | Rural | 8063 (61.2%) | 3210 (59.7%) |
|  | Urban | 5120 (38.8%) | 2170 (40.3%) |
|  | Poorest | 2420 (18.4%) | 877 (16.3%) |
|  | Poorer | 2363 (17.9%) | 1040 (19.3%) |
|  | Middle | 2372 (18%) | 957 (17.8%) |
|  | Richer | 2810 (21.3%) | 1047 (19.5%) |
|  | Richest | 3218 (24.4%) | 1458 (27.1%) |
| **Education level** | No education | 3522 (26.7%) | 581 (10.8%) |
|  | Basic | 5601 (42.5%) | 2556 (47.5%) |
|  | Secondary or higher | 4060 (30.8%) | 2243 (41.7%) |
| **Employment** | No | 9178 (69.6%) | 1013 (18.8%) |
|  | Yes | 4005 (30.4%) | 4367 (81.2%) |
| **Marital status** | Married/cohabit | 8488 (64.4%) | 3132 (58.2%) |
|  | Separated/divorced | 1799 (13.6%) | 270 (5.0%) |
|  | Single | 2896 (22.0%) | 1979 (36.8%) |
| **Health** | Bad | 145 (1.1%) | 280 (5.2%) |
|  | Good | 9975 (75.7%) | 4137 (76.9%) |
|  | Moderate | 3063 (23.2%) | 963 (17.9%) |

**Table 1.** Survey weighted prevalence estimates of social demographic characteristics of the study participants.

| **Climate variable** | **Source** | **Spatial resolution (Native)** | **Temporal resolution (Native)** |
| --- | --- | --- | --- |
| Annual daytime temperature ( ^o^C ) | ERA5-Land (ECMWF)-2m temperature | 1 ^o^ (~9km) | Hourly |
| Annual daytime relative humidity (%) | ERA5-Land (ECMWF)-derived from 2m temperature and dew point | 1 ^o^ (~9km) | Hourly |
| Annual mean nighttime temperature ( ^o^C ) | ERA5-Land (ECMWF)-2m temperature | 1 ^o^ (~9km) | Hourly |
| Annual nighttime relative humidity (%) | ERA5-Land (ECMWF)- derived from 2m temperature and dewpoint | 1 ^o^ (~9km) | Hourly |
| Annual mean daily relative humidity (%) | ERA5-Land (ECMWF)- derived from 2m temperature and dewpoint | 1 ^o^ (~9km) | Hourly |
| Annual mean daily temperature  ( ^o^C ) | ERA5-Land (ECMWF)-2m temperature | 1 ^o^ (~9km) | Hourly |
| Normalized Difference Vegetation Index (NDVI) | Modis Terra Vegetation Indices (MODIS13A2, collection 6) | 1km | 16-day composite |
| Total annual Precipitation | CHIRPS | (~5.6km) | Monthly |
| Universal Thermal Climate Index (UTCI) | GLoUTCI | 1km | Monthly |

**Table 2.** Table showing source and characteristics of climatic variables used in the study.

**Environmental Exposure definitions**

(a) **Temperature**

- Nighttime exposure was defined as 21:00 to 05: 00 reflecting common initiation sleeping times ^1,2^ and daytime exposure as 06:00 to 18:00, reflecting typical hours of outdoor activities and day-time heat exposure. Daily exposure period covered a continuous 24-hour period.
- Annual mean daily temperature was calculated by averaging hourly 2m temperature values over 24-hour period, which were then averaged over 12 months preceding the 2022-23 Mozambique DHS. Similarly, annual day time and nighttime temperature measurements were derived using daytime and nighttime exposure definitions, respectively.

(b) **Relative humidity**

- Relative humidity was computed using the Magnus method which utilizes 2m air temperature (T) and dewpoint temperature (T_d_)^3,4^.

Saturation vapor:

$$e_{s}\left( T \right)=6.112e^{\frac{17.67T}{T+243.5}}$$

Actual vapor pressure:

$$e\left( T_{d} \right)=6.112e^{\frac{17.67Td}{Td+243.5}}$$

And relative humidity (RH):

$$RH(\%)=\frac{100e\left( T_{d} \right)}{e_{s}\left( T \right)}$$

- Using the above formula, annual daily relative humidity was then estimate using hourly 2m air temperature and 2m dewpoint temperature measurements over 24-hour period, which was then averaged over 12 months preceding the 2022-23 Mozambique DHS. In a similarly manner, annual day time and nighttime relative humidity were derived using day time and nighttime exposure periods.

**Universal Thermal Climate Index**

- The universal thermal climate index (UTCI) is a bioclimatic measure of describing the physiological comfort of the human body under specific weather conditions^5^.
- This composite measure derived from ambient temperature, relative humidity, wind and radiation indicates thermal stress levels, with values above 46 representing extreme heat stress and those value below -40 indicating extreme cold stress^5^.
- Monthly UTCI values were obtained from the Global Universal Thermal Climate Index (GLoUTCI) database and averaged over 12 months preceding the 2022-23 MDHS.

**Precipitation**

- Total annual precipitation (which includes all forms of water that fall from the atmosphere such as rain, drizzle or hail) was obtained by aggregating monthly precipitation estimates from the Climate Hazards Group InfraRed Precipitation (CHIRPS) station data to represent cumulative rainfall exposure over the 12 months preceding the survey^6^.

**Normalized Difference Vegetation Index**

- Vegetation health using the Normalized Difference Vegetation Index (NDVI) was derived from MODIS 16-day composite product (MODI13A2, collection 6)^7^. This index ranging from -1 to 1 quantifies vegetation vigor and density based on the difference between near infrared (NIR) and red (RED) reflectance expressed as:

$$NDVI=\frac{NIR-RED}{NIR+RED}$$

- We averaged 23 composites (16-day composites) spanning from January to December of 2021.

| **Climatic/ environmental exposure variable** | **Min** | **Max** | **Median (IQR)** | **Mean (SD)** |
| --- | --- | --- | --- | --- |
| NDVI | 0.21 | 0.75 | 0.51 (0.42-0.55) | 0.49 (0.1) |
| Precipitation (mm) | 159 | 1,999 | 1074 (794-1287) | 1067 (308) |
| Daytime temperature (^o^C) | 20.1 | 28.1 | 25.7 (24.0-26.7) | 25.4 (1.6) |
| Daytime relative humidity (%) | 47.1 | 73.6 | 63.4 (60.7-66.6) | 63.4 (4.3) |
| Nighttime temperature (^o^C) | 16.6 | 24.7 | 21.9 (20.9-22.8) | 21.7 (1.7) |
| Nighttime relative humidity (%) | 58.7 | 88.8 | 80.9 (78.1-83.7) | 80.5 (4.9) |
| Daily temperature (^o^C) | 18.4 | 26.3 | 23.8 (22.5-24.9) | 23.6 (1.6) |
| Daily relative humidity (%) | 52.6 | 78.5 | 72.3 (69.3-74.3) | 71.6 (4.2) |
| UTCI | 12.1 | 34.5 | 30.1 (28.2-31.5) | 29.5 (2.9) |

**Table 3**. Summary statistics of climatic and environmental variables used in the study

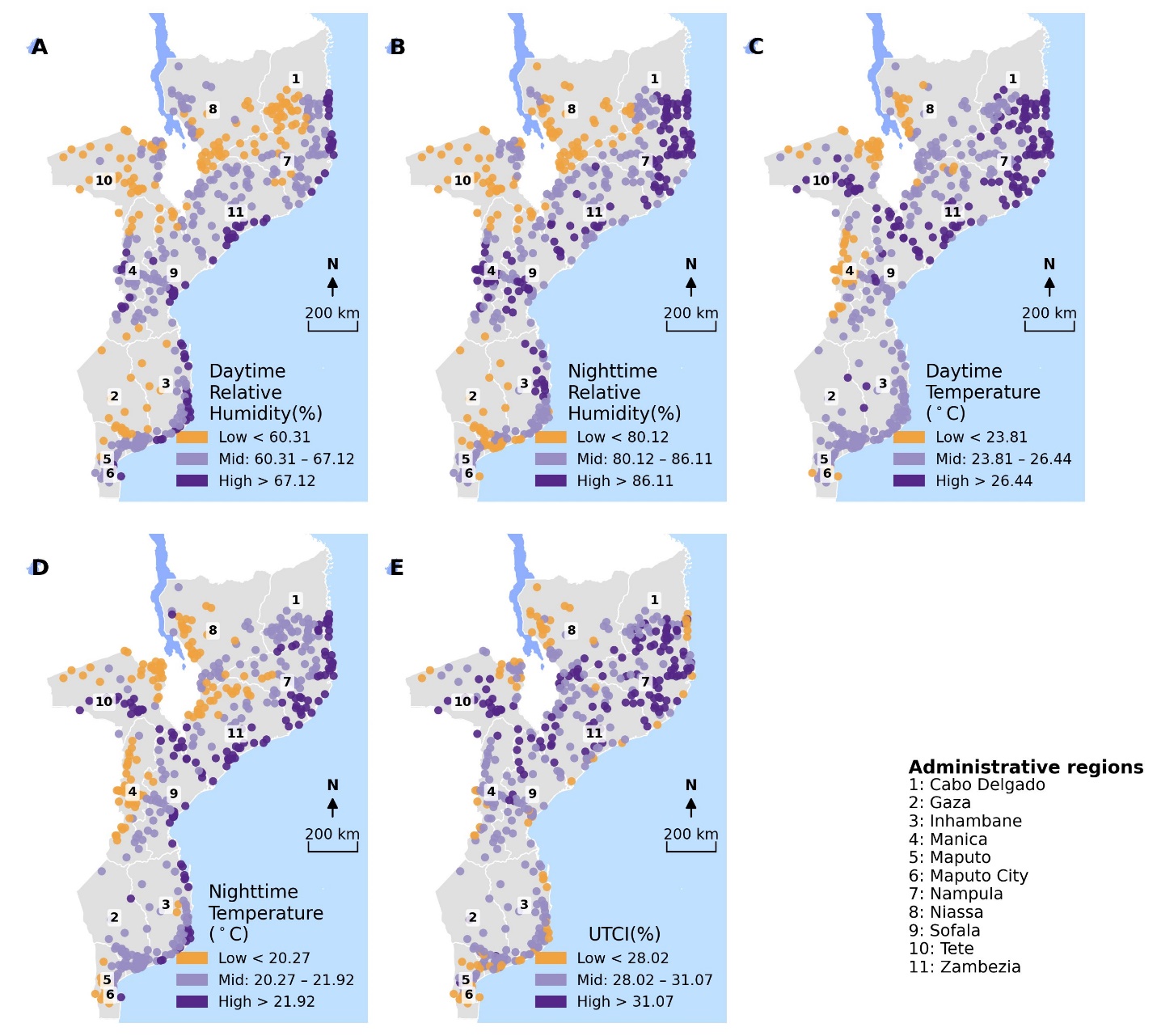

**Fig 1**. Spatial distribution of environmental factors at DHS survey locations. (**A**): Daytime relative humidity, (**B**): Nighttime relative humidity, (**C**): Daytime temperature, (**D**): Nighttime temperature, (**E**): Universal Thermal Climate Index (UCTI).

| **Variable** | **Exposure prevalence** | | **Anxiety** | | **Depression** | | **Anxiety and depression** | |
| --- | --- | --- | --- | --- | --- | --- | --- | --- |
|  | **Level** | **No. (%)** | **No. (%)** | **P_trend** | **No. (%)** | **P_trend** | **No. (%)** | **P_trend** |
| **Daily temp.** | Low | 3288 ( 24.9 ) | 171 ( 5.2 ) | 0.003 | 125 ( 3.8 ) | 0.001 | 83 ( 2.5 ) | 0.004 |
|  | Moderate | 5711 ( 43.3 ) | 517 ( 9.1 ) |  | 482 ( 8.4 ) |  | 345 ( 6.0 ) |  |
|  | High | 4184 ( 31.7 ) | 775 ( 18.5 ) |  | 730 ( 17.4 ) |  | 538 ( 12.9 ) |  |
| **Precip.** | Low | 3021 ( 22.9 ) | 172 ( 5.7 ) | 0.017 | 123 ( 4.1 ) | 0.013 | 89 ( 2.9 ) | 0.019 |
|  | Moderate | 5262 ( 39.9 ) | 504 ( 9.6 ) |  | 509 ( 9.7 ) |  | 328 ( 6.2 ) |  |
|  | High | 4901 ( 37.2 ) | 786 ( 16.0 ) |  | 704 ( 14.4 ) |  | 549 ( 11.2 ) |  |
| **Daily humidity** | Low | 2759 ( 20.9 ) | 144 ( 5.2 ) | 0.072 | 129 ( 4.7 ) | 0.059 | 68 ( 2.5 ) | 0.058 |
|  | Moderate | 6992 ( 53.0 ) | 874 ( 12.5 ) |  | 778 ( 11.1 ) |  | 583 ( 8.3 ) |  |
|  | High | 3431 ( 26.0 ) | 443 ( 12.9 ) |  | 430 ( 12.5 ) |  | 315 ( 9.2 ) |  |
| **NDVI** | Low | 3406 ( 25.8 ) | 415 ( 12.2 ) | 0.204 | 357 ( 10.5 ) | 0.552 | 272 ( 8.0 ) | 0.456 |
|  | Moderate | 6761 ( 51.3 ) | 847 ( 12.5 ) |  | 739 ( 10.9 ) |  | 534 ( 7.9 ) |  |
|  | High | 3016 ( 22.9 ) | 201 ( 6.7 ) |  | 241 ( 8.0 ) |  | 160 ( 5.3 ) |  |
| **Daily temp.** | High | 1766 ( 32.8 ) | 38 ( 2.2 ) | 0.8842 | 42 ( 2.4 ) | 0.9243 | 12 ( 0.7 ) | 0.9308 |
|  | Low | 1344 ( 25.0 ) | 32 ( 2.4 ) |  | 29 ( 2.2 ) |  | 10 ( 0.7 ) |  |
|  | Moderate | 2270 ( 42.2 ) | 43 ( 1.9 ) |  | 51 ( 2.2 ) |  | 14 ( 0.6 ) |  |
| **PRECIP** | High | 2079 ( 38.6 ) | 35 ( 1.7 ) | 0.7690 | 41 ( 2.0 ) | 0.9629 | 13 ( 0.6 ) | 0.9324 |
|  | Low | 1193 ( 22.2 ) | 29 ( 2.4 ) |  | 36 ( 3.0 ) |  | 9 ( 0.8 ) |  |
|  | Moderate | 2108 ( 39.2 ) | 48 ( 2.3 ) |  | 45 ( 2.1 ) |  | 15 ( 0.7 ) |  |
| **Daily relative humidity** | High | 1418 ( 26.4 ) | 36 ( 2.5 ) | 0.7039 | 25 ( 1.8 ) | 0.8524 | 11 ( 0.8 ) | 0.8155 |
|  | Low | 1125 ( 20.9 ) | 29 ( 2.6 ) |  | 35 ( 3.1 ) |  | 13 ( 1.2 ) |  |
|  | Moderate | 2837 ( 52.7 ) | 48 ( 1.7 ) |  | 62 ( 2.2 ) |  | 13 ( 0.5 ) |  |
| **NDVI** | High | 1178 ( 21.9 ) | 29 ( 2.5 ) | 0.7376 | 23 ( 2.0 ) | 0.8898 | 4 ( 0.3 ) | 0.8620 |
|  | Low | 1444 ( 26.8 ) | 35 ( 2.4 ) |  | 51 ( 3.5 ) |  | 17 ( 1.2 ) |  |
|  | Moderate | 2758 ( 51.3 ) | 49 ( 1.8 ) |  | 48 ( 1.7 ) |  | 15 ( 0.5 ) |  |

**Table 4.** Survey weighted prevalence estimates of symptoms of generalized anxiety, depression, and co-occurring symptoms of anxiety and depression by environmental exposures.

| **Disorder** | **Variable** | **F_value** | **Num df** | **Den df** | **P value** |
| --- | --- | --- | --- | --- | --- |
| **Anxiety** | NDVI | 8.71 | 2 | 588 | <0.0001 |
|  | Precipitation | 8.56 | 2 | 588 | <0.0001 |
|  | Relative humidity | 9.69 | 2 | 588 | <0.0001 |
|  | Temperature | 25.82 | 2 | 588 | <0.0001 |
| **Depression** | NDVI | 3.71 | 2 | 588 | 0.0252 |
|  | Precipitation | 6.62 | 2 | 588 | 0.0014 |
|  | Relative humidity | 8.01 | 2 | 588 | < 0.0001 |
|  | Temperature | 27.98 | 2 | 588 | < 0.0001 |
| **Anxiety and depression** | NDVI | 3.83 | 2 | 588 | 0.0222 |
|  | Precipitation | 5.11 | 2 | 588 | 0.0063 |
|  | Relative humidity | 5.63 | 2 | 588 | 0.0038 |
|  | Temperature | 18.27 | 2 | 588 | <0.0001 |

**Table 5**. Wald F-test results for the overall associations (adjusted for socio-demographic factors) between environmental exposure variables and mental health outcomes among women.

| **Variable** | **Anxiety** | | **Depression** | | **Anxiety and depression** | |
| --- | --- | --- | --- | --- | --- | --- |
|  | aRR (95% CI) | P Value | aRR (95% CI) | P Value | aRR (95% CI) | P Value |
| **Residence (ref=rural)** | | | | | | |
| Urban | 0.93 (0.64–1.35) | 0.6984 | 1.07 (0.76–1.49) | 0.7043 | 1.05 (0.67–1.65) | 0.8195 |
| **Education(ref=none)** |  |  |  |  |  |  |
| Basic | 1.15 (0.96–1.39) | 0.1397 | 1.11 (0.91–1.34) | 0.2936 | 1.24 (0.97–1.58) | 0.0827 |
| Secondary or higher | 1.21 (0.94–1.55) | 0.1449 | 0.96 (0.73–1.26) | 0.7691 | 1.09 (0.78–1.53) | 0.6063 |
| **Wealth index(ref=poorest)** | | | | | | |
| Poorer | 0.78 (0.60–1.01) | 0.0611 | 0.81 (0.61–1.08) | 0.1519 | 0.78 (0.57–1.07) | 0.1240 |
| Middle | 0.76 (0.57–1.01) | 0.0601 | 0.72 (0.54–0.98) | 0.0343 | 0.55 (0.37–0.82) | 0.0038 |
| Richer | 0.75 (0.56–1.02) | 0.0660 | 0.74 (0.55–1.01) | 0.0553 | 0.72 (0.49–1.07) | 0.1026 |
| Richest | 0.51 (0.35–0.76) | 0.0009 | 0.52 (0.36–0.77) | 0.0011 | 0.42 (0.25–0.69) | 0.0007 |
| **Marital status(ref=married/cohabiting)** | | | | | | |
| Separated/divorced | 1.27 (1.07–1.51) | 0.0070 | 1.28 (1.06–1.54) | 0.0105 | 1.42 (1.13–1.78) | 0.0026 |
| Single | 0.86 (0.64–1.17) | 0.3344 | 1.06 (0.80–1.41) | 0.6731 | 0.96 (0.65–1.40) | 0.8215 |
| **Employment(ref=no)** | | | | | | |
| Yes | 0.80 (0.66–0.96) | 0.0176 | 0.78 (0.65–0.94) | 0.0086 | 0.72 (0.57–0.92) | 0.0088 |
| **Age(ref=15-19 years)** | | | | | | |
| 20-24 | 1.24 (1.00–1.52) | 0.0463 | 1.28 (1.02–1.62) | 0.0367 | 1.31 (1.01–1.70) | 0.0448 |
| 25-29 | 1.25 (0.96–1.62) | 0.1004 | 1.18 (0.91–1.54) | 0.2194 | 1.14 (0.81–1.59) | 0.4570 |
| 30-34 | 1.20 (0.89–1.62) | 0.2289 | 1.12 (0.85–1.48) | 0.4263 | 1.09 (0.77–1.56) | 0.6180 |
| 35-39 | 1.60 (1.22–2.08) | 0.0006 | 1.34 (1.04–1.74) | 0.0244 | 1.49 (1.08–2.04) | 0.0142 |
| 40-44 | 1.12 (0.82–1.52) | 0.4765 | 1.01 (0.74–1.38) | 0.9466 | 0.96 (0.66–1.41) | 0.8529 |
| 45-49 | 1.36 (1.04–1.79) | 0.0275 | 1.26 (0.95–1.68) | 0.1121 | 1.33 (0.95–1.87) | 0.0979 |
| **Self-reported health status(ref=bad)** | | | | | | |
| Moderate | 0.92 (0.60–1.40) | 0.6827 | 1.07 (0.66–1.73) | 0.7905 | 1.11 (0.62–1.98) | 0.7216 |
| Good | 0.60 (0.40–0.91) | 0.0172 | 0.61 (0.37–1.01) | 0.0564 | 0.62 (0.35–1.09) | 0.0992 |
| **Normalized difference vegetation index (ref=Low)** | | | | | | |
| Moderate | 0.67 (0.46–0.98) | 0.0388 | 0.71 (0.49–1.03) | 0.0708 | 0.65 (0.40–1.04) | 0.0722 |
| High | 0.34 (0.20–0.57) | <0.0001 | 0.51 (0.31–0.83) | 0.0068 | 0.42 (0.22–0.77) | 0.0058 |
| **Precipitation (ref=Low)** |  |  |  |  |  |  |
| Moderate | 1.28 (0.88–1.86) | 0.1984 | 1.63 (1.09–2.46) | 0.0189 | 1.33 (0.77–2.27) | 0.3060 |
| High | 1.98 (1.35–2.92) | 0.0005 | 2.20 (1.41–3.41) | 0.0005 | 2.09 (1.20–3.64) | 0.0098 |
| **Relative humidity (ref=Low)** | | | | | | |
| Moderate | 2.21 (1.52–3.21) | <0.0001 | 2.08 (1.41–3.07) | 0.0003 | 2.85 (1.49–5.45) | 0.0016 |
| High | 1.54 (1.00–2.38) | 0.0513 | 1.51 (0.97–2.36) | 0.0691 | 2.09 (1.04–4.19) | 0.0389 |
| **Temperature (ref=Low)** | | | | | | |
| Moderate | 1.45 (1.06–1.97) | 0.0196 | 1.79 (1.26–2.56) | 0.0012 | 1.85 (1.17–2.93) | 0.0091 |
| High | 2.89 (2.13–3.93) | <0.0001 | 3.45 (2.46–4.84) | <0.0001 | 3.64 (2.35–5.64) | <0.0001 |

**Table 6**. Estimates of adjusted risk ratios of associations between mental health outcomes and environmental, and socio-demographic factors among women. The categories low (<25the percentile), moderate (25th-75th percentile) and high (>75the percentile) reflect increasing exposure burden for environmental factors.

| **Variable** | **Anxiety** | | **Depression** | | **Anxiety and depression** | |
| --- | --- | --- | --- | --- | --- | --- |
|  | aRR (95% CI) | P Value | aRR (95% CI) | P Value | aRR (95% CI) | P Value |
| **Residence (ref=rural)** | | | | | | |
| Urban | 2.45 (1.28–4.69) | 0.0070 | 2.83 (1.55–5.17) | 0.0007 | 6.48 (1.29–32.56) | 0.0236 |
| **Education(ref=none)** |  |  |  |  |  |  |
| Basic | 1.81 (0.37–8.86) | 0.4631 | 0.96 (0.36–2.53) | 0.9330 | 6.41 (0.82–50.36) | 0.0778 |
| Secondary or higher | 2.85 (0.58–13.92) | 0.1967 | 1.43 (0.44–4.71) | 0.5555 | 7.01 (0.92–53.41) | 0.0605 |
| **Wealth index(ref=poorest)** | | | | | | |
| Poorer | 1.06 (0.40–2.87) | 0.9010 | 0.69 (0.27–1.80) | 0.4491 | 2.65 (0.40–17.52) | 0.3110 |
| Middle | 0.69 (0.26–1.88) | 0.4704 | 1.06 (0.28–4.00) | 0.9353 | 2.56 (0.63–10.46) | 0.1917 |
| Richer | 0.80 (0.30–2.11) | 0.6521 | 0.69 (0.25–1.94) | 0.4862 | 1.62 (0.31–8.41) | 0.5631 |
| Richest | 0.57 (0.19–1.75) | 0.3284 | 0.94 (0.28–3.13) | 0.9161 | 1.49 (0.30–7.40) | 0.6267 |
| **Marital status(ref=married/cohabiting)** | | | | | | |
| Separated/divorced | 2.84 (1.43–5.62) | 0.0029 | 2.84 (1.58–5.13) | 0.0005 | 3.76 (1.55–9.13) | 0.0035 |
| Single | 1.58 (0.67–3.72) | 0.2999 | 1.43 (0.56–3.66) | 0.4503 | 1.54 (0.66–3.62) | 0.3180 |
| **Employment(ref=no)** | | | | | | |
| Yes | 1.28 (0.70–2.34) | 0.4258 | 0.52 (0.33–0.84) | 0.0078 | 0.48 (0.20–1.17) | 0.1072 |
| **Age(ref=15-19 years)** | | | | | | |
| 20-24 | 0.90 (0.40–2.06) | 0.8112 | 0.67 (0.25–1.74) | 0.4079 | 0.58 (0.05–6.17) | 0.6528 |
| 25-29 | 1.44 (0.55–3.75) | 0.4584 | 1.87 (0.80–4.36) | 0.1475 | 3.67 (1.09–12.37) | 0.0367 |
| 30-34 | 1.33 (0.44–4.05) | 0.6149 | 1.51 (0.59–3.89) | 0.3929 | 2.71 (0.91–8.09) | 0.0752 |
| 35-39 | 3.28 (0.94–11.43) | 0.0621 | 2.96 (1.18–7.40) | 0.0208 | 7.20 (1.75–29.64) | 0.0064 |
| 40-44 | 1.57 (0.44–5.61) | 0.4915 | 2.23 (0.66–7.54) | 0.1976 | 3.34 (0.68–16.50) | 0.1397 |
| 45-49 | 1.54 (0.48–4.95) | 0.4732 | 1.91 (0.53–6.86) | 0.3193 | 1.24 (0.12–13.08) | 0.8588 |
| 50-54 | 2.69 (0.77–9.36) | 0.1212 | 2.45 (0.70–8.58) | 0.1608 | 4.46 (0.83–23.98) | 0.0819 |
| **Self-reported health status(ref=bad)** | | | | | | |
| Moderate | 0.38 (0.17–0.83) | 0.0158 | 0.43 (0.21–0.91) | 0.0269 | 0.43 (0.13–1.36) | 0.1504 |
| Good | 0.72 (0.30–1.73) | 0.4639 | 0.65 (0.31–1.38) | 0.2651 | 0.19 (0.05–0.72) | 0.0148 |
| **Normalized difference vegetation index (ref=Low)** | | | | | | |
| Moderate | 2.62 (1.13–6.06) | 0.0247 | 1.67 (0.74–3.76) | 0.2180 | 1.04 (0.26–4.11) | 0.9562 |
| High | 1.31 (0.65–2.64) | 0.4528 | 0.96 (0.56–1.65) | 0.8905 | 0.90 (0.36–2.25) | 0.8146 |
| **Precipitation (ref=Low)** |  |  |  |  |  |  |
| Moderate | 0.79 (0.38–1.62) | 0.5150 | 1.26 (0.63–2.51) | 0.5067 | 1.93 (0.58–6.43) | 0.2836 |
| High | 0.84 (0.45–1.58) | 0.5968 | 1.07 (0.62–1.87) | 0.8015 | 1.25 (0.42–3.75) | 0.6844 |
| **Relative humidity (ref=Low)** | | | | | | |
| Moderate | 0.75 (0.37–1.52) | 0.4229 | 0.32 (0.15–0.68) | 0.0033 | 0.34 (0.12–1.01) | 0.0522 |
| High | 0.47 (0.25–0.89) | 0.0216 | 0.48 (0.29–0.79) | 0.0046 | 0.23 (0.08–0.62) | 0.0041 |
| **Temperature (ref=Low)** | | | | | | |
| Moderate | 0.88 (0.44–1.75) | 0.7109 | 1.42 (0.79–2.57) | 0.2427 | 0.75 (0.21–2.71) | 0.6643 |
| High | 0.84 (0.50–1.42) | 0.5121 | 1.07 (0.59–1.95) | 0.8176 | 0.79 (0.35–1.79) | 0.5706 |

**Table 7**. Estimates of adjusted risk ratios of associations between common mental health outcomes and environmental, and socio-demographic factors among men. The categories low (<25the percentile), moderate (25th-75th percentile) and high (>75the percentile) reflect increasing exposure burden for environmental factors.

|  | **Exposure**  **Level** | **Anxiety** | | **Depression** | | **Anxiety and depression** | |
| --- | --- | --- | --- | --- | --- | --- | --- |
|  |  | **aRR (95% CI)** | **P value** | **aRR (95% CI)** | **P value** | **aRR (95% CI)** | **P value** |
| **Women** | | | | | | | |
| **Daytime temp(ref=Low)** | Moderate | 2.32 (1.74–3.10) | <0.0001 | 2.42 (1.75–3.35) | <0.0001 | 3.22 (2.13–4.86) | <0.0001 |
|  | High | 4.45 (3.28–6.05) | <0.0001 | 4.56 (3.29–6.32) | <0.0001 | 6.43 (4.24–9.76) | <0.0001 |
| **Daytime rel. humid(ref=Low)** | Moderate | 1.58 (1.09–2.29) | 0.0169 | 1.44 (0.96–2.16) | 0.0804 | 1.65 (0.90–3.00) | 0.1037 |
|  | High | 1.01 (0.67–1.53) | 0.9573 | 0.99 (0.61–1.58) | 0.9525 | 1.04 (0.54–2.00) | 0.9155 |
| **Precipitation**  **(ref=Low)** | Moderate | 1.07 (0.69–1.65) | 0.7762 | 1.50 (0.94–2.40) | 0.0905 | 1.15 (0.61–2.17) | 0.6743 |
|  | High | 1.74 (1.13–2.69) | 0.0124 | 2.09 (1.29–3.38) | 0.0030 | 1.90 (1.01–3.58) | 0.0462 |
| **Night-time temperature (ref=Low)** | Moderate | 1.40 (1.00–1.97) | 0.0505 | 2.04 (1.40–2.96) | <0.0001 | 1.85 (1.12–3.05) | 0.0161 |
|  | High | 1.73 (1.24–2.42) | 0.0014 | 2.33 (1.60–3.40) | <0.0001 | 2.17 (1.35–3.48) | 0.0014 |
| **Night time rel. humidity (ref=Low)** | Moderate | 1.84 (1.28–2.63) | 0.0011 | 1.74 (1.17–2.58) | 0.0061 | 2.27 (1.23–4.19) | 0.0090 |
|  | High | 3.04 (2.01–4.59) | <0.0001 | 2.70 (1.77–4.10) | <0.0001 | 4.06 (2.09–7.89) | <0.0001 |
| **Men** | | | | | | | |
| **Daytime temp(ref=Low)** | Moderate | 0.85 (0.51–1.42) | 0.5411 | 0.73 (0.44–1.20) | 0.2092 | 0.63 (0.26–1.48) | 0.2878 |
|  | High | 0.93 (0.47–1.81) | 0.8234 | 1.33 (0.70–2.51) | 0.3882 | 0.57 (0.15–2.19) | 0.4129 |
| **Daytime rel. humid(ref=Low)** | Moderate | 0.75 (0.42–1.36) | 0.3510 | 0.70 (0.40–1.21) | 0.2029 | 0.43 (0.17–1.11) | 0.0813 |
|  | High | 0.86 (0.39–1.93) | 0.7216 | 0.48 (0.24–0.95) | 0.0361 | 0.59 (0.22–1.57) | 0.2906 |
| **Night-time temperature (ref=Low)** | Moderate | 0.70 (0.40–1.25) | 0.2287 | 1.18 (0.58–2.40) | 0.6404 | 0.82 (0.37–1.84) | 0.6326 |
|  | High | 0.75 (0.39–1.44) | 0.3844 | 1.22 (0.68–2.21) | 0.5000 | 0.55 (0.17–1.74) | 0.3105 |
| **Night time rel. humidity (ref=Low)** | Moderate | 0.39 (0.20–0.75) | 0.0046 | 0.37 (0.21–0.64) | 0.0005 | 0.21 (0.07–0.58) | 0.0029 |
|  | High | 0.72 (0.39–1.34) | 0.2983 | 0.32 (0.16–0.64) | 0.0012 | 0.41 (0.15–1.14) | 0.0890 |

**Table 4**. Estimates of adjusted risk ratios of the associations between daytime and nighttime temperature and relative humidity exposures, and common mental disorders among women and men. In addition to socio-demographic factors, the models were adjusted for NVDI and total annual precipitation. The categories low (<25the percentile), moderate (25th-75th percentile) and high (>75the percentile) reflect increasing exposure burden for environmental factors.

| **Exposure** | **Exposure**  **Level** | **Anxiety** | | **Depression** | | **Anxiety and depression** | |
| --- | --- | --- | --- | --- | --- | --- | --- |
|  |  | **aRR (95% CI)** | **P value** | **aRR (95% CI)** | **P value** | **aRR (95% CI)** | **P value** |
| **Women** | | | | | | | |
| **UTCI (ref=Low)** | Moderate | 1.00 (0.66–1.50) | 0.9850 | 1.13 (0.73–1.75) | 0.5923 | 1.15 (0.66–2.02) | 0.6239 |
|  | High | 1.90 (1.29–2.79) | 0.0011 | 2.17 (1.42–3.31) | <0.0001 | 2.28 (1.36–3.84) | 0.0020 |
| **Men** | | | | | | | |
| **UTCI (ref=Low)** | Moderate | 1.24 (0.66–2.35) | 0.5058 | 0.89 (0.51–1.57) | 0.6861 | 0.64 (0.24–1.75) | 0.3885 |
|  | High | 0.97 (0.40–2.36) | 0.9424 | 1.16 (0.53–2.52) | 0.7072 | 0.61 (0.17–2.24) | 0.4579 |

**Table 8**. Estimates of adjusted risk ratios of the associations between Universal Thermal Climate Index (UTCI) and other environmental factors and mental disorders among women and men. The models were adjusted for NVDI, precipitation and socio demographic factors. The categories low (<25the percentile), moderate (25th-75th percentile) and high (>75the percentile) reflect increasing exposure burden for environmental factors.

| **Mental disorder** | **Climatic Exposure** | **Exposure Level** | **15-22 Years** | | **23-32 Years** | | **33-49 Years** | |
| --- | --- | --- | --- | --- | --- | --- | --- | --- |
|  |  |  | aRR (95% CI) | P  Value | aRR (95% CI) | P  value | aRR (95% CI) | P  value |
| **Symptoms of**  **anxiety** | Daily Temperature | Moderate | 1.69 (1.04–2.74) | 0.035 | 1.21 (0.75–1.93) | 0.433 | 1.5 (1.04–2.16) | 0.030 |
|  |  | High | 3.77 (2.37–6.00) | <0.001 | 2.56 (1.66–3.96) | <0.001 | 2.5 (1.67–3.72) | <0.001 |
|  | NDVI | Moderate | 0.63 (0.38–1.03) | 0.063 | 0.78 (0.50–1.20) | 0.258 | 0.62 (0.41–0.94) | 0.024 |
|  |  | High | 0.38 (0.20–0.72) | 0.004 | 0.37 (0.19–0.70) | 0.003 | 0.28 (0.16–0.51) | <0.001 |
|  | Precipitation | Moderate | 0.89 (0.54–1.45) | 0.629 | 1.51 (0.86–2.66) | 0.150 | 1.38 (0.88–2.16) | 0.160 |
|  |  | High | 1.47 (0.86–2.52) | 0.158 | 1.76 (0.98–3.16) | 0.059 | 2.76 (1.76–4.31) | <0.001 |
|  | Daily Relative Humidity | Moderate | 2.33 (1.35–4.03) | 0.002 | 2.27 (1.40–3.68) | <0.001 | 2.17 (1.45–3.25) | <0.001 |
|  |  | High | 1.61 (0.89–2.93) | 0.117 | 1.56 (0.91–2.67) | 0.106 | 1.50 (0.86–2.60) | 0.154 |
| **Symptoms of**  **depression** | Daily Temperature | Moderate | 2.05 (1.18–3.53) | 0.011 | 1.71 (1.05–2.77) | 0.030 | 1.66 (1.04–2.63) | 0.033 |
|  |  | High | 4.94 (2.96–8.25) | <0.001 | 2.89 (1.79–4.68) | <0.001 | 2.74 (1.70–4.42) | <0.001 |
|  | NDVI | Moderate | 0.79 (0.48–1.29) | 0.343 | 0.70 (0.45–1.09) | 0.120 | 0.62 (0.40–0.97) | 0.037 |
|  |  | High | 0.75 (0.40–1.39) | 0.365 | 0.42 (0.23–0.77) | 0.005 | 0.39 (0.21–0.71) | 0.002 |
|  | Precipitation | Moderate | 1.98 (1.11–3.53) | 0.020 | 1.56 (0.90–2.72) | 0.112 | 1.35 (0.86–2.13) | 0.193 |
|  |  | High | 2.69 (1.44–5.04) | 0.002 | 1.63 (0.93–2.86) | 0.092 | 2.38 (1.44–3.94) | <0.001 |
|  | Daily Relative Humidity | Moderate | 1.76 (1.02–3.04) | 0.044 | 2.54 (1.57–4.11) | <0.001 | 2.12 (1.34–3.34) | 0.001 |
|  |  | High | 1.06 (0.57–1.97) | 0.847 | 2.15 (1.22–3.78) | 0.008 | 1.51 (0.86–2.67) | 0.155 |
| **Co-occurring symptoms of**  **anxiety**  **and depression** | Daily Temperature | Moderate | 1.77 (0.91–3.45) | 0.095 | 1.72 (0.91–3.23) | 0.094 | 1.96 (1.09–3.53) | 0.026 |
|  |  | High | 3.96 (2.13–7.39) | <0.001 | 3.68 (2.01–6.75) | <0.001 | 2.99 (1.59–5.62) | <0.001 |
|  | NDVI | Moderate | 0.72 (0.38–1.37) | 0.320 | 0.64 (0.38–1.08) | 0.098 | 0.56 (0.31–1.00) | 0.050 |
|  |  | High | 0.51 (0.23–1.16) | 0.110 | 0.37 (0.18–0.75) | 0.006 | 0.36 (0.17–0.77) | 0.009 |
|  | Precipitation | Moderate | 1.22 (0.58–2.57) | 0.603 | 1.55 (0.73–3.30) | 0.255 | 1.13 (0.63–2.04) | 0.682 |
|  |  | High | 2.12 (0.96–4.67) | 0.062 | 1.73 (0.81–3.71) | 0.157 | 2.44 (1.30–4.57) | 0.005 |
|  | Daily Relative Humidity | Moderate | 2.55 (1.05–6.18) | 0.039 | 3.28 (1.54–7.00) | 0.002 | 2.93 (1.49–5.72) | 0.002 |
|  |  | High | 1.77 (0.70–4.51) | 0.228 | 2.40 (1.04–5.50) | 0.040 | 2.12 (0.97–4.64) | 0.059 |

**Table 9**. Estimates of adjusted risk ratios of the associations between environmental factors and mental disorders among women by age group. All models were adjusted for education, health status, urbanity, household wealth index and marital status. The categories low (<25the percentile), moderate (25th-75th percentile) and high (>75the percentile) reflect increasing exposure burden for environmental factors.

| **Exposure** | **Anxiety** | | **Depression** | | **Anxiety and depression** | |
| --- | --- | --- | --- | --- | --- | --- |
|  | **aRR (95% CI)** | **P value** | **aRR (95% CI)** | **P value** | **aRR (95% CI)** | **P value** |
| **Women** | | | | | | |
| **NVDI ( per 0.1)** | 0.68 (0.58–0.81) | <0.0001 | 0.80 (0.66–0.97) | 0.0209 | 0.74 (0.60–0.91) | 0.0040 |
| **Precipitation (per 100mm)** | 1.11 (1.06–1.16) | <0.0001 | 1.10 (1.05–1.15) | <0.0001 | 1.12 (1.05–1.19) | <0.0001 |
| **Daily temperature (^o^C)** | 1.30 (1.21–1.40) | <0.0001 | 1.35 (1.24–1.47) | <0.0001 | 1.38 (1.24–1.52) | <0.0001 |
| **Daily relative humidity** | 1.04 (1.01–1.06) | 0.0080 | 1.04 (1.01–1.07) | 0.0067 | 1.05 (1.01–1.09) | 0.0111 |
| **Men** | | | | | | |
| **NVDI ( per 0.1)** | 1.43 (1.05–1.96) | 0.0258 | 0.99 (0.72–1.36) | 0.9375 | 0.71 (0.44–1.14) | 0.1561 |
| **Precipitation (per 100mm)** | 0.95 (0.87–1.03) | 0.2389 | 1.05 (0.96–1.14) | 0.3302 | 1.07 (0.93–1.24) | 0.3396 |
| **Daily temperature (^o^C)** | 1.02 (0.85–1.24) | 0.8146 | 1.04 (0.88–1.23) | 0.6442 | 0.93 (0.68–1.25) | 0.6143 |
| **Daily relative humidity** | 0.98 (0.92–1.05) | 0.6050 | 0.92 (0.87–0.97) | 0.0024 | 0.96 (0.88–1.05) | 0.3699 |

**Table 10**. Estimates of adjusted risk ratios of the associations between continuous environmental factors and mental disorders among women and men. The models were adjusted for age, education, health status, urbanity, marital status, employment and household wealth index.
